# Network anatomy in logopenic variant of primary progressive aphasia

**DOI:** 10.1101/2023.05.15.23289065

**Authors:** Maria Luisa Mandelli, Diego L. Lorca-Puls, Sladjana Lukic, Maxime Montembeault, Andrea Gajardo-Vidal, Abigail Licata, Aaron Scheffler, Giovanni Battistella, Stephanie M Grasso, Rian Bogley, Buddhika M. Ratnasiri, Renaud La Joie, Nidhi S. Mundada, Eduardo Europa, Gil Rabinovici, Bruce L. Miller, Jessica De Leon, Maya L. Henry, Zachary Miller, Maria Luisa Gorno-Tempini

## Abstract

The logopenic variant of primary progressive aphasia (lvPPA) is a neurodegenerative syndrome characterized linguistically by gradual loss of repetition and naming skills, resulting from left posterior temporal and inferior parietal atrophy. Here, we sought to identify which specific cortical loci are initially targeted by the disease (epicenters) and investigate whether atrophy spreads through pre-determined networks. First, we used cross-sectional structural MRI data from individuals with lvPPA to define putative disease epicenters using a surface-based approach paired with an anatomically-fine-grained parcellation of the cortical surface (i.e., HCP-MMP1.0 atlas). Second, we combined cross-sectional functional MRI data from healthy controls and longitudinal structural MRI data from individuals with lvPPA to derive the epicenter-seeded resting-state networks most relevant to lvPPA symptomatology and ascertain whether functional connectivity in these networks predicts longitudinal atrophy spread in lvPPA. Our results show that two partially distinct brain networks anchored to the left anterior angular and posterior superior temporal gyri epicenters were preferentially associated with sentence repetition and naming skills in lvPPA. Critically, the strength of connectivity within these two networks in the neurologically-intact brain significantly predicted longitudinal atrophy progression in lvPPA. Taken together, our findings indicate that atrophy progression in lvPPA, starting from inferior parietal and temporo-parietal junction regions, predominantly follows at least two partially non-overlapping pathways, which may influence the heterogeneity in clinical presentation and prognosis.

## 1. Introduction

Several studies of selective network vulnerability in neurodegenerative diseases have shown that misfolded proteins propagate trans-neuronally in a prion-like manner from most vulnerable regions (i.e., disease epicenter) to connected structures (Prusiner 1984; Zhou et al. 2012; Liu et al. 2012; Iba et al. 2013; Ahmed et al. 2014; Goedert, Eisenberg, and Crowther 2017). This network-based neurodegeneration hypothesis has received support from animal studies (Clavaguera et al. 2009; de Calignon et al. 2012; Iba et al. 2013), as well as from an increasing number of human neuroimaging studies demonstrating that neurodegeneration spreads along pre-determined large-scale neural networks of functionally and structurally connected regions that closely mirror the patterns of atrophy seen in distinct neurodegenerative syndromes (Seeley et al. 2009; Zhou et al. 2010; Mandelli et al. 2016; Cope et al. 2018; Franzmeier and Dyrba 2017; Vogel et al. 2020).

Primary progressive aphasia (PPA) is a family of neurodegenerative syndromes characterized by a relatively isolated and progressive loss of speech and language abilities that occurs when either frontotemporal lobar degeneration (FTLD) or Alzheimer’s disease (AD) pathology preferentially targets language-related brain networks. Within this family, three main variants have been described (Gorno-Tempini et al. 2011): 1) the non-fluent/agrammatic variant (nfvPPA), characterized by impaired motor speech and/or agrammatism, and most often associated with abnormal deposition of microtubule-associated protein tau (Josephs et al. 2006; Kertesz et al. 2005; Spinelli et al. 2017); 2) the semantic variant (svPPA), characterized by impaired naming and single word comprehension, and most often associated with abnormal deposition of transactive response DNA-binding protein of 43kD (TDP-43) (Davies et al. 2005; Snowden, Neary, and Mann 2007; Marsel Mesulam et al. 2008; Spinelli et al. 2017); and 3) the logopenic variant (lvPPA), characterized by impaired single-word retrieval in spontaneous speech and naming and impaired repetition of phrases and sentences, and most often caused by underlying AD pathology (Mesulam 2008; Rabinovici et al. 2008; Migliaccio et al. 2009; Grossman 2010; Rohrer, Rossor, and Warren 2012; Kirshner 2012; Harris and Jones 2014; Spinelli et al. 2017; Giannini et al. 2017).

In line with the network-based neurodegeneration hypothesis, current neuroimaging evidence indicates that the loss of brain tissue (i.e., atrophy) in nfvPPA originates in the left posterior inferior frontal gyrus and spreads within the speech production network to the precentral gyrus and supplementary motor area, supramarginal gyrus, posterior temporal regions, and subcortical and insular regions (Mandelli et al. 2016). Damage to these regions has been shown to be associated with motor speech and syntactic impairments (Wilson, Dronkers, et al. 2010; Mandelli et al. 2018; 2014; García et al. 2022). In svPPA, atrophy-related changes start in the left anterior temporal lobe and spread within the semantic network to the ventral and medial temporal lobe, cingulate, insular, orbitofrontal cortex, and angular gyrus (Collins et al. 2017; Brown et al. 2019; Brambati et al. 2007; Rosen et al. 2002). Damage to these regions has been previously associated with word comprehension and naming deficits (Chow et al. 2010; Migliaccio et al. 2016; Cousins et al. 2016; Joubert et al. 2017). On the other hand, in lvPPA, a few studies have revealed substantial variability regarding the location and extent of early atrophy, which often includes a large region comprising the left posterior temporal and inferior parietal cortices known to be susceptible to tau deposition and neurodegeneration in AD (Gorno-Tempini et al. 2008; R. Migliaccio et al. 2009; Rohrer et al. 2013; Whitwell et al. 2019; Rik Ossenkoppele et al. 2016; Conca et al. 2022). Crucially, the lack of a well-defined epicenter in lvPPA hinders the mapping of specific networks related to lvPPA symptomatology and disease progression. This inconsistency across prior studies might at least in part be related to differences in methodology (volumetric studies or studies addressing cortical thickness or metabolic changes), the choice of anatomical parcellation atlases, and different nomenclature adopted by different investigators to describe brain regions in and around the left temporo-parietal junction (Gorno-Tempini et al. 2008; Migliaccio et al. 2009; Rohrer et al. 2013; Rogalski et al. 2016; Phillips et al. 2018). Moreover, recent studies show that atrophy in lvPPA-related regions may underlie non-linguistic symptoms in lvPPA, such as deficits in attention and visuospatial functions (Ramanan et al. 2022), leaving open the question of which specific epicenters and associated networks are relevant for naming and repetition deficits, the two core aphasic symptoms in lvPPA. Therefore, the precise localization of early atrophy and its progression remains to be established. This information is also critical for tracking longitudinal changes in clinical trials and specifying potential target brain regions for brain stimulation in therapeutic trials (tDCS and TMS) (Gervits et al. 2016; Ficek et al. 2018; Tsapkini et al. 2018; Nissim, Moberg, and Hamilton 2020).

In this study, we performed a cross-sectional and longitudinal multimodal MRI investigation in a large cohort of well-characterized individuals with lvPPA and healthy controls. By leveraging an anatomically-fine-grained atlas, we sought to improve the localization of early changes in cortical thickness in lvPPA, map the functional networks anchored to these regions in healthy controls, and test whether the strength of functional connectivity in language-relevant networks derived from the neurologically-intact brain predict longitudinal atrophy spread in lvPPA.

## 2. Material and Methods

### 2.1 Participants

From the database of the Memory and Aging Center at the University of California, San Francisco (UCSF-MAC), we selected individuals who had (i) an imaging-supported diagnosis of lvPPA according to published international criteria (Gorno-Tempini et al. 2011), (ii) a high-resolution structural 3D-T1 MRI scan that passed quality check, and (iii) a Mini-Mental State Examination (MMSE) score ≥ 15 and a Clinical Dementia Rating (CDR) < 2. A total of 91 individuals with lvPPA were identified, and among them, 48 had a PET scan with ^11^C-Pittsburgh Compound B (PIB, for amyloid-β pathology) and 22 had autopsy-proven pathology (of which 8 had a PIB-PET scan). One out of 48 had a negative PIB-PET scan, while four out of 22 had mixed or non-AD pathology. These participants were excluded from the study. Within the total cohort, we then isolated two non-overlapping groups that had either a positive PIB-PET scan or autopsy-proven AD pathology as follows: (1) a group for the cross-sectional analysis (lvPPA_mild_, n=15; all of them had a positive PIB-PET scan and one had, in addition, autopsy-proven AD pathology) with a milder clinical presentation (MMSE > 20 and CDR < 1); (2) a group for the longitudinal analysis (lvPPA_long,_ n=28; 24 had a positive PIB-PET scan; 13 had autopsy-proven AD pathology; 7 had both) with two MRI scans at least 1 year apart.

Three distinct groups of cognitively and neurologically intact individuals were also selected: (1) a group for the cross-sectional analysis (HC_cross_, n=68) matched to the lvPPA_tot_ group for age, sex, handedness, education, and scanner type; (2) a group for the longitudinal analysis (HC_long_, n=56) matched to the lvPPA_long_ group for age, sex, handedness, education, and scanner type; and (3) a group of individuals (HC_conn_, n=50) with resting state functional MRI (rsfMRI) data for the delineation of the healthy intrinsic connectivity networks (ICNs).

All individuals with lvPPA underwent a comprehensive evaluation including neurological history and examination, neuropsychological assessment (Kramer et al. 2003), and speech and language testing (Gorno-Tempini et al. 2004; Wilson, Henry, et al. 2010). Written informed consent was obtained from all participants or their surrogates. The study was approved by the University of California (San Francisco and Berkeley) and Lawrence Berkeley National Laboratory (LBNL) institutional review boards for human research.

### 2.2 Image acquisition and processing

#### 2.2.1 PET Imaging

Amyloid PET scans were acquired using ^11^C-Pittsburgh compound B at LBNL on either a Siemens ECAT Exact HR PET only scanner (n=18) or a Siemens Biograph 6 Truepoint PET/CT scanner (n=30), as previously described (Villeneuve et al. 2015). PIB was synthesized and radiolabeled at LBNL’s Biomedical Isotope Facility. Attenuation correction was performed using an emission scan (ECAT scanner) or a low-dose CT scan (Biograph scanner) acquired prior to PET acquisition. PET data were acquired in list mode and reconstructed using an ordered subset expectation maximization algorithm with weighted attenuation and smoothed with a 4 mm Gaussian kernel with scatter correction. PIB Standardized Uptake Value Ratio (SUVR) images were based on mean uptake over 50-70 min post-injection of ∼15mCi of PIB (four 5-min frames).

PET frames were realigned, averaged and co-registered onto their corresponding structural MRI. PIB SUVR images were created in native space using the MRI-defined cerebellar gray matter as a reference region (Villeneuve et al. 2015). PIB-PET positivity was based on both visual reading, as validated against autopsy findings (La Joie et al. 2019; Lesman-Segev et al. 2020), and a quantitative analysis of the PIB-PET data. Briefly, the average PIB-SUVR is extracted from a large neocortical region of interest (ROI); resulting values are considered positive when exceeding a threshold of 1.21 (Villeneuve et al. 2015).

#### 2.2.2 Structural MRI

All brain structural MRI scans were acquired with either a 1.5T Vision, 3T Trio or 3T Prisma scanner (Siemens Healthcare). For the longitudinal scans, we only included those collected using the same scanner at both time points. All the structural images were acquired with a T1-weighted 3D magnetization prepared rapid acquisition gradient echo (MPRAGE) sequence with the following parameters: 164 coronal slices; voxel size = 1.0 x 1.5 x 1.0 mm^3^; FoV = 256×256 mm^2^; matrix size = 256×256; TR = 10 msec; TE = 4 msec; T1= 300 msec; flip angle = 15° (1.5T); 160 sagittal slices; voxel size = 1.0 x 1.0 x 1.0 mm^3^; FoV = 256×256 mm^2^; matrix size = 256×256; TR = 2300 msec; TE = 2.98 msec for 3T Trio and TE = 2.9 msec for 3T Prisma; flip angle = 9° (3T).

All T1-weighted images were first visually assessed to ensure the absence of artifacts or excessive motion. The images were processed through the Computational Anatomy Toolbox (CAT12; http://dbm.neuro.uni-jena.de/cat, version 12.7) with the Statistical Parametric Mapping software (SPM12; http://www.fil.ion.ucl.ac.uk/spm/software/spm12) running under Matlab 2020b (http://www.mathworks.com).

The whole process consists of three steps: 1) an initial voxel-based processing, 2) a main voxel-based processing, and 3) a surface-based processing. In step (1) the images undergo a spatial adaptive non-local means (SANLM) denoising filter (Manjón et al. 2010). After bias correction and affine registration, the data undergo standard SPM unified segmentation (Ashburner and Friston 2005). In step (2), the output images were skull-stripped and the brain parcellated into left and right hemisphere, subcortical regions and cerebellum. All the segmented tissue classes underwent a local intensity transformation before the final adaptive maximum a posteriori (AMAP) segmentation (Rajapakse, Giedd, and Rapoport 1997), which is refined by applying a partial volume (Tohka, Zijdenbos, and Evans 2004). Finally, the segmented images were spatially normalized to a common reference space using the geodesic shooting registration (Ashburner and Friston 2011). For the longitudinal data, the images were realigned from the two time points using inverse-consistent rigid-body registrations and intra-subject bias field corrections were applied (Ashburner and Ridgway 2012; Reuter and Fischl 2011; Reuter et al. 2012), thus assuring comparability across the time points. For the spatial registration to the reference brain template, a mean transformation for the time points was calculated and applied to all individual images. The use of an unbiased average image reduces random variations in the processing procedure and improves the robustness and sensitivity of the longitudinal analysis. In step (3), cortical thickness estimation and reconstruction of the central surface took place using a projection-based thickness method (Dahnke, Yotter, and Gaser 2013). After the initial surface reconstruction, topological defects were repaired using spherical harmonics (Yotter et al. 2010), and the final result is a central surface mesh that provides the basis for extracting folding patterns where the resulting local values were projected onto each node. Finally, the individual central surfaces were spatially registered to an average template using a spherical mapping (Yotter et al. 2011). Cortical thickness values were extracted using the HCP-MMP1.0 (Human Connectome Project Multi-Modal Parcellation version 1.0) parcellation. This atlas was generated using multi-modal brain images and an objective semi-automated neuroanatomical approach under the Human Connectome Project (HCP) (http://www.humanconnectomeproject.org/) and provides a fine-grained parcellation of the human brain into a total of 360 regions (180 per hemisphere). For a detailed description of parcellation nomenclature, we refer the reader to the neuroanatomical supplementary information provided in Glasser et al. (Glasser et al. 2016).

#### 2.2.3 Functional imaging processing

RsfMRI data were acquired on the 3T Prisma scanner equipped with a 64-channel head coil and using a T2*-weighted multiband-EPI pulse sequences including 560 volumes with 66 AC/PC-aligned axial slices in interleaved order (slice thickness = 2.2 mm; in-plane resolution = 2.2×2.2 mm^2^; FoV = 211×211 mm^2^; TR = 850 msec; TE = 32.80 msec; flip angle = 45°; multiband factor of 6). The participants were asked to lie still in the scanner with eyes closed, remain awake and think of nothing. RsfMRI data were visually checked for quality control and only participants with a maximum relative head motion less than 2mm for translation and 3 degrees for rotation were included. A total of 50 healthy controls matched by age and sex to the lvPPA group were included in the study (HC_conn_, n=50). Data were analyzed using an in-house pipeline combining different tools from FSL (http://fsl.fmrib.ox.ac.uk/fsl/fslwiki/), AFNI (http://afni.nimh.nih.gov/afni/), SPM (https://www.fil.ion.ucl.ac.uk/spm/software/spm12/), and ANTs (https://www.nitrc.org/projects/ants). The pre-processing steps involved slice timing correction and realignment of the functional whole-brain volumes, after discarding the first 5 volumes. Susceptibility induced distortions characteristic of EPI acquisitions were estimated and corrected using the TOPUP tool (https://fsl.fmrib.ox.ac.uk/fsl/fslwiki/topup), which makes use of two additional spin-echo images acquired with opposing polarities of the phase-encode blips. The mean functional image was subsequently normalized to the MNI template using a combination of rigid, linear, and nonlinear warping, as implemented in ANTs. The ICA-AROMA tool was then utilized to identify and remove motion-related components from rsfMRI data in a data-driven fashion by means of an independent component analysis (ICA) (Pruim et al. 2015). Finally, time-series were band-pass filtered and CSF and WM mean signals were regressed out from the data and finally smoothed with an isotropic Gaussian kernel of 5mm.

### 2.3 Statistical Analyses

The analyses aim to identify which of the anatomically parcellated regions are involved in mild lvPPA (epicenters), map the functional networks anchored to these regions in healthy controls, and identify which of the language-relevant networks are related to disease progression in lvPPA.

In this section, we describe in detail the steps used to (1) convert cortical thickness values in each region of the atlas to the W-score scale, (2) identify the regions of peak atrophy in the mild cohort of lvPPA, (3) derive the ICNs in the healthy individuals using the most atrophic regions from the analysis performed in 2.3.2 as seeds, (4) determine the ICNs most relevant to lvPPA symptomatology, (5) assess the cortical thickness change for each regions in the longitudinal cohort of lvPPA, and (6) test if the behaviorally relevant networks were significant predictors of atrophy progression in lvPPA. Finally, additional statistical analyses of behavioral data were performed using independent and paired t-tests on the measures of dementia severity and language performance available in the participants both cross-sectionally and longitudinally.

#### 2.3.1 Conversion of cortical thickness values to W-score scale

For each participant, mean cortical thickness values were extracted and covariate-adjusted on an ROI-by-ROI basis using the brain parcellation of the HCP-MMP1.0 atlas. Specifically, the influence of age, sex, handedness, and scanner type on cortical thickness was covaried out by fitting a multiple regression model to the data from the group of healthy controls (HC_cross_). Covariate-adjusted cortical thickness values (i.e., W-scores) were calculated for each ROI as follows: (observed cortical thickness – expected cortical thickness) / standard deviation of the residuals for that ROI in the healthy controls.

#### 2.3.2 Definition of the most vulnerable regions in the mild lvPPA cohort

Cross-sectional cortical thickness values on the W-score scale were modeled using ROI-specific simple linear regression models with the main effect of diagnostic status to identify the regions with the most significant cortical atrophy in the mild cohort of lvPPA (lvPPA_mild_, n=15) compared to the matched group of healthy controls (HC_cross,_ n=68). Structural and metabolic studies have consistently shown that the left inferior parietal and temporo-parietal junction are regions particularly vulnerable to neurodegeneration in lvPPA (Conca et al., 2022). Therefore, we predicted that a significant disease epicenter (i.e., a cluster of atrophic regions) would be identified within the left parieto-temporal cortex. In addition, since the size of the patient cohort used to identify the most atrophic regions was small (n=15), we repeated the same analysis using the baseline MRI scans from the longitudinal and total cohorts of lvPPA patients (n=28 and n=86, respectively) to assess the reproducibility and generalizability of the results.

#### 2.3.3 Intrinsic connectivity network maps in healthy controls

The ICN maps were obtained by performing a seed-to-voxel analysis across the whole brain in the HC_conn_ (n=50) group using the regions with the most significant difference between groups (lvPPA_mild_ n=15 versus HC_cross_ n=68) as seeds. For each of these seed regions, the average time series was extracted and used to compute the temporal correlation against all other voxels with a Pearson’s r coefficient. Correlation coefficients were then converted to Z-scores by Fisher’s r-to-Z transformation and group-level functional connectivity maps were derived by performing one-sample *t*-tests in SPM12 with the null hypothesis set to zero and only positive correlations being tested. In order to enhance the robustness of the results and minimize the risk of false positives, we adopted a more stringent statistical threshold of *P* < 0.001, family wise error (FWE) corrected. Binary masks of each thresholded functional network were then created. For each functional network, regions from the HCP-MMP1.0 parcellation with at least 70% of overlap with the corresponding binary mask were considered part of that network.

#### 2.3.4 Identification of language-relevant ICNs in lvPPA

In order to ascertain the behavioral relevance of the ICNs in relation to the core language deficits in lvPPA, we performed a correlation analysis (involving the full cohort of 86 individuals with lvPPA) between atrophy (average W-score) within each of the networks and severity of the two defining clinical symptoms of lvPPA: impaired sentence repetition and confrontation naming (Gorno-Tempini et al., 2011). Specifically, we calculated the mean cortical thickness within each ICN in the total cohort of lvPPA (n=86) as the average of W-scores (see 2.3.1) across all regions belonging to that ICN (see 2.3.3). We then derived one behavioral index of sentence repetition ability and another one of confrontation naming ability based on the six most difficult sentences from the Western Aphasia Battery Repetition subtest (WAB, Kertesz 1980) and Boston Naming Test (BNT; Kaplan, Goodglass, and Weintraub 1983), respectively. Repetition and naming are both tasks that involve language production. Therefore, in order to isolate task components preferentially related to auditory-verbal short-term memory (in sentence repetition) or lexical retrieval (in confrontation naming), we conducted partial correlation analyses (focusing on one variable while accounting for the other). Based on previous studies, we hypothesized that regions anchored to the temporo-parietal junction would show stronger association with sentence repetition (Hickok and Poeppel 2004), while the network connected to one or more subregions of the angular gyrus would be preferentially involved in lexical retrieval (Battistella et al. 2020).

#### 2.3.5 Identification of regions that demonstrate longitudinal atrophy-related changes in lvPPA

Cortical thickness values for each ROI in each participant with longitudinal data (lvPPA_long_ n=28) were converted into W-scores as described above in Section 2.3.1. Longitudinal trajectories of cortical thickness values on the W-score scale were modeled using ROI-specific linear mixed models (LMM) with main effects for diagnostic status, time (defined as months since first scan), and the two-way interaction of diagnostic status and time. Subject-specific random intercepts were included to account for dependencies due to repeated observations within participants. For each ROI, the interaction term between diagnostic status and time captures differences in the longitudinal trajectory of cortical atrophy between diagnostic groups over the study period. The corresponding t-statistic for the interaction term was treated as an index of the difference in cortical thickness change between the two diagnostic groups over time for each ROI defined in the parcellation atlas.

#### 2.3.6 Assessment of atrophy progression within the ICNs most relevant to the core language symptoms in lvPPA

By resorting to graph theoretical analysis, we calculated the shortest functional path from every ROI of the atlas parcellation to the seed-ROI of each ICN that showed an association with sentence repetition or naming deficits, which are the two defining symptoms of lvPPA (see 2.3.4). We then used this metric (shortest functional path obtained from the healthy control group, n=50) as a predictor of a region’s degree of longitudinal atrophy progression in individuals with lvPPA (n=28) (i.e., t-statistic from 2.3.5) as previously reported (Zhou et al. 2012; Mandelli et al., 2016; Brown et al. 2019). The inputs to the graph theory analysis were the regions of the HCP-MMP1.0 atlas (as nodes) and the Z-scores between pair of regions (as edges). The shortest functional path was calculated for each ROI as the sum of the edge weights forming the shortest path from a given ROI to the seed (Rubinov and Sporns 2010) using an unthresholded connectivity matrix in the Brain Connectivity Toolbox (BCT, https://www.nitrc.org/projects/bct). Following the network-based neurodegeneration hypothesis (Seeley et al. 2009; Zhou et al. 2010; Mandelli et al. 2016; Brown et al. 2019), a shorter path to the epicenter was expected to be associated with greater longitudinal cortical change. Multiple regression analyses were performed on the t-statistic for the interaction term of the cortical change for each ROI between the two diagnostic groups over time (see Section 2.3.5) and the shortest path length to the seed-ROI. The Euclidean distance between the center of mass of each ROI and the epicenter of each ICN of interest was also calculated and included in the statistical analysis to account for the spatial proximity between each region and the seed.

We chose the salience network as a control network, as it is not usually involved in lvPPA. The salience network is composed of the anterior insula, dorsal anterior cingulate cortex, and frontoparietal operculum, and has been previously implicated in detecting and filtering important sensory, emotional, and cognitive information (Seeley et al. 2007). The salience network was seeded from the right anterior insula (right anterior agranular insula complex in the HCP-MMP1.0 atlas) using the same cohort of healthy controls (n=50).

#### 2.3.7 Behavioral statistical analyses

Independent t-tests and paired t-tests were performed in the cross-sectional and longitudinal groups of lvPPA for MMSE, CDR Sum of Boxes, and language tasks to detect differences from the cognitively normal performance and longitudinal changes over the study period, respectively. Independent t-tests were also performed between the mild group of lvPPA patients and the remaining lvPPA patients (n=71) in order to highlight differences in symptom severity (see Table 1). When departures from normality of the data were detected, non-parametric alternatives were adopted.

**Table 1.**
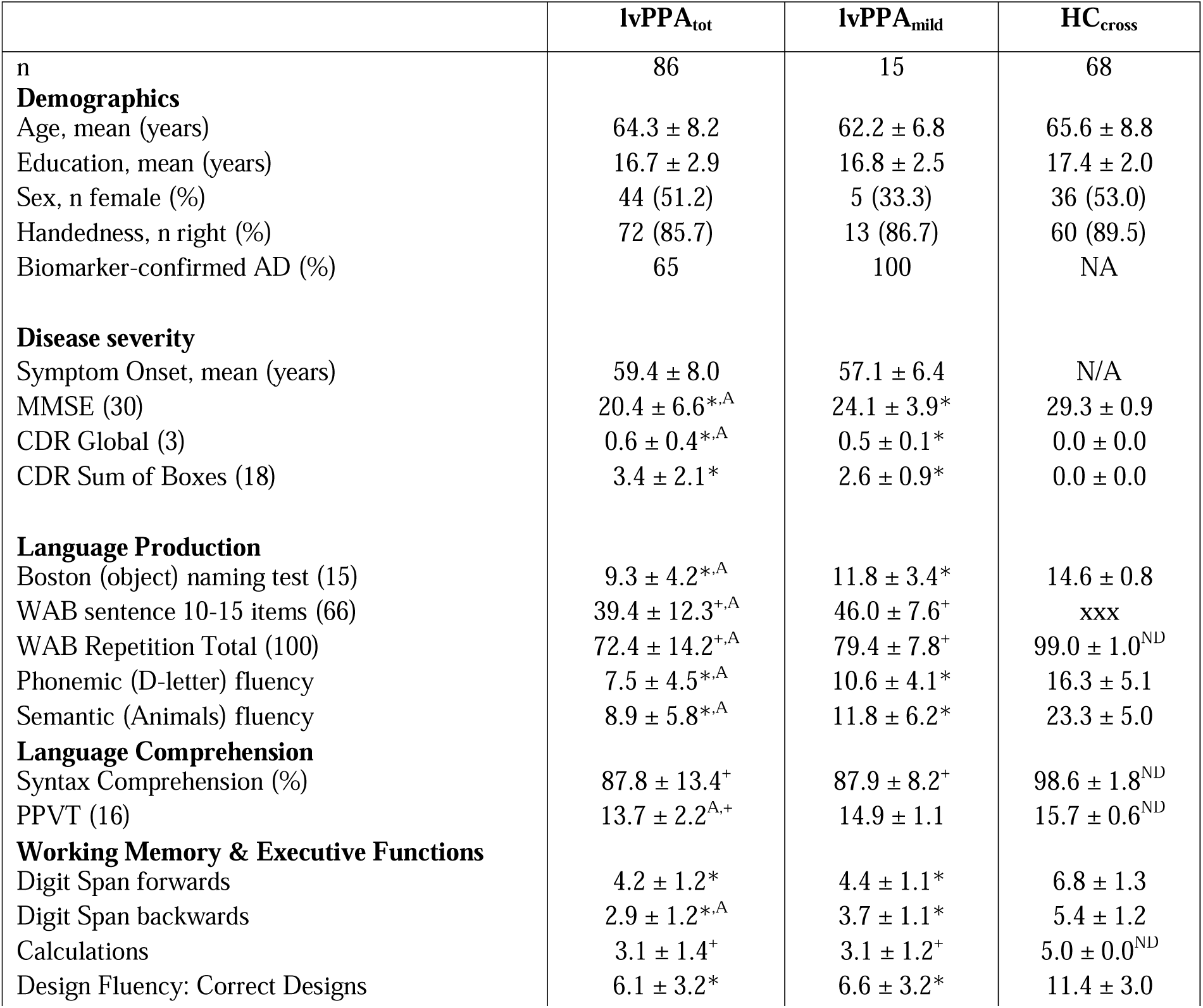
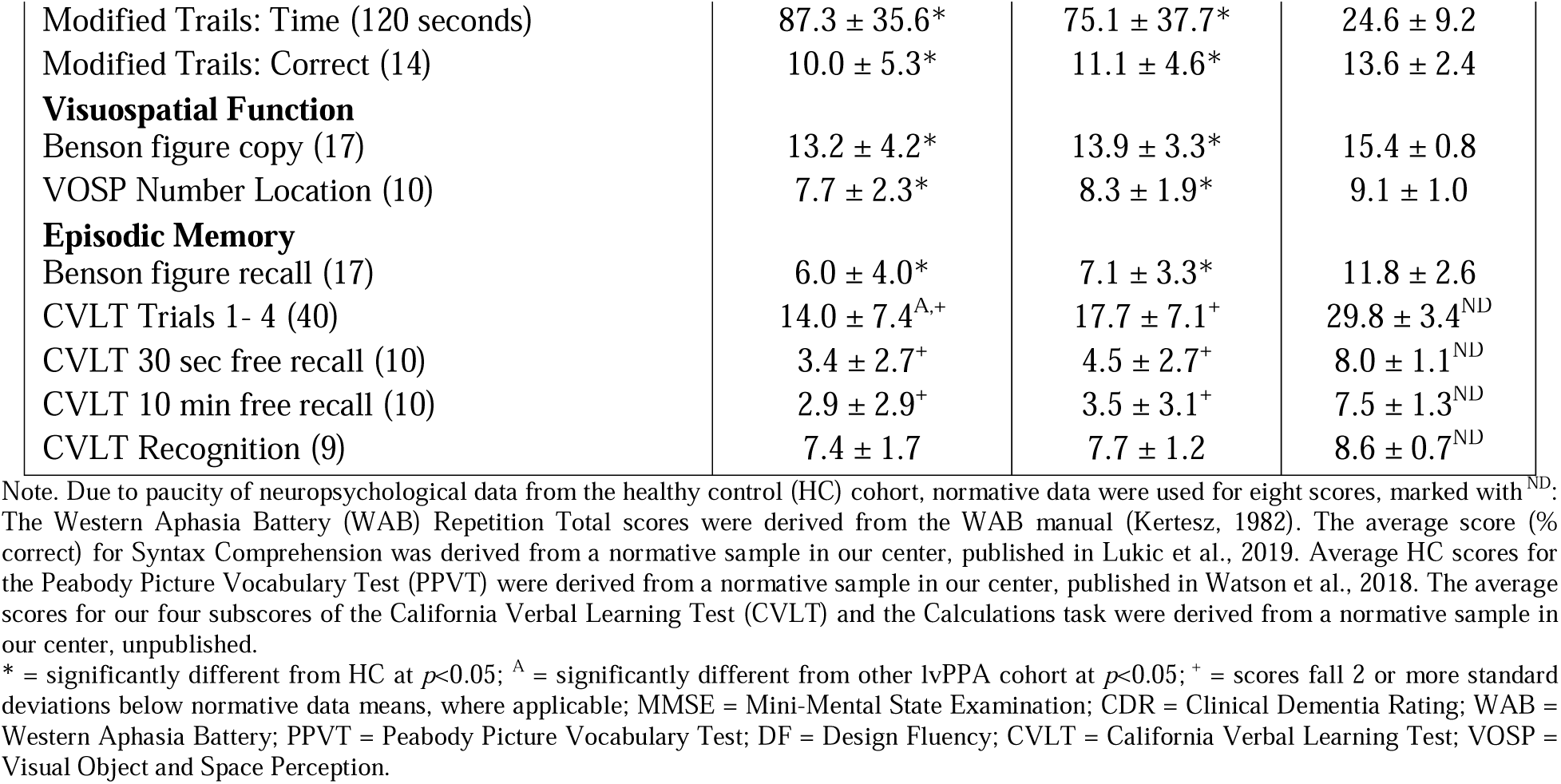
Summary of demographic and cognitive profiles for the individuals with lvPPA and healthy controls used for the cross-sectional analysis.

### 2.4 Data statement

While we are able to share anonymized data, public archiving is not yet permitted under the study’s IRB approval due to the sensitive nature of the data. Specific requests can be submitted through the UCSF-MAC Resource (Request form: http://memory.ucsf.edu/resources/data). Following a UCSF-regulated procedure, access will be granted to designated individuals in line with ethical guidelines on the reuse of sensitive data. This would require submission of a Material Transfer Agreement, available at: https://icd.ucsf.edu/material-transfer-and-data-agreements.

## 3. Results

### 3.1 Participants and clinical observation

Demographic and clinical data of the total cohort of individuals with lvPPA (lvPPA_tot,_ *n*=86) are summarized in Table 1. This table also includes a description of the subgroup of individuals with mild symptoms (lvPPA_mild_, *n*=15) as well as the healthy control group used for the cross-sectional analysis (HC_cross_, *n*=68). Both the total cohort of lvPPA and the mild group exhibited the typical profile of impairment in verbal short-term memory (WAB repetition, digit span forward and California Verbal Learning Test (CVLT) trials 1-4) and confrontation naming when compared to healthy controls. Relative to the lvPPA_mild_ group, the lvPPA_tot_ group generally showed more severe impairment in overall cognition (lower MMSE and higher CDR scores) as well as language production (BNT, repetition, phonemic fluency), CVLT trials 1-4, and digit span backward. The total cohort of lvPPA encompasses the entire spectrum of impairment including the mild group. The statistical comparisons are indicated in Table 1.

Table 2 provides summary demographic and clinical information for the two cohorts with longitudinal structural MRI data (lvPPA_long_, *n*=28; HC_long_, *n*=56). Paired t-tests performed in the lvPPA_long_ group showed longitudinal worsening of overall cognition (MMSE/CDR), core lvPPA symptoms (verbal short-term memory and naming) and decline of additional language skills (verbal fluency, single-word comprehension and sentence comprehension). With the exception of verbal episodic memory and arithmetic abilities, other cognitive domains, such as visual episodic memory, executive functioning, visuospatial and visuo-constructive abilities, did not decline significantly between the two time points. This indicates that the progression of cognitive decline between the two time points within this cohort of patients was mostly restricted to the language domain, while other cognitive domains remained relatively stable across time.

**Table 2.**
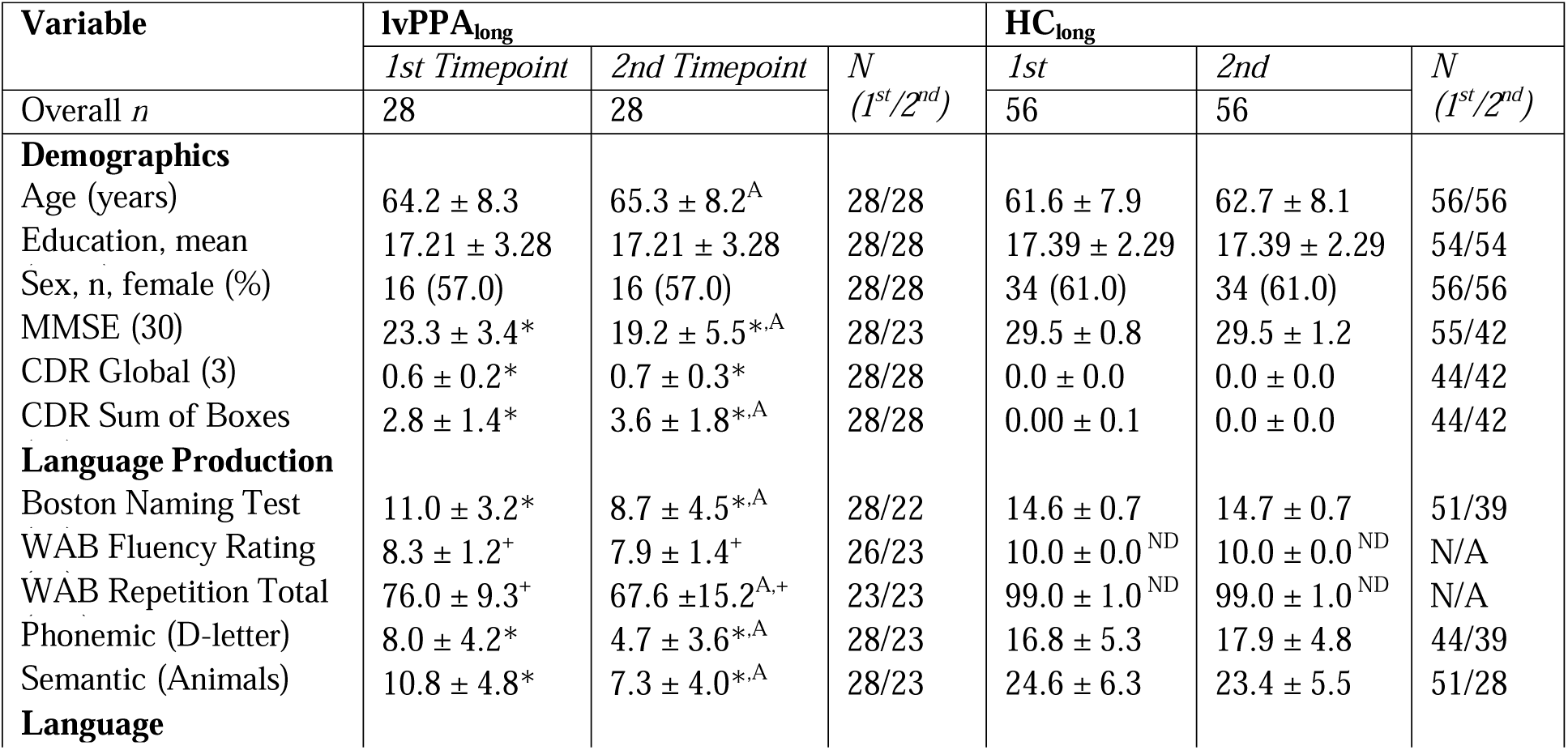
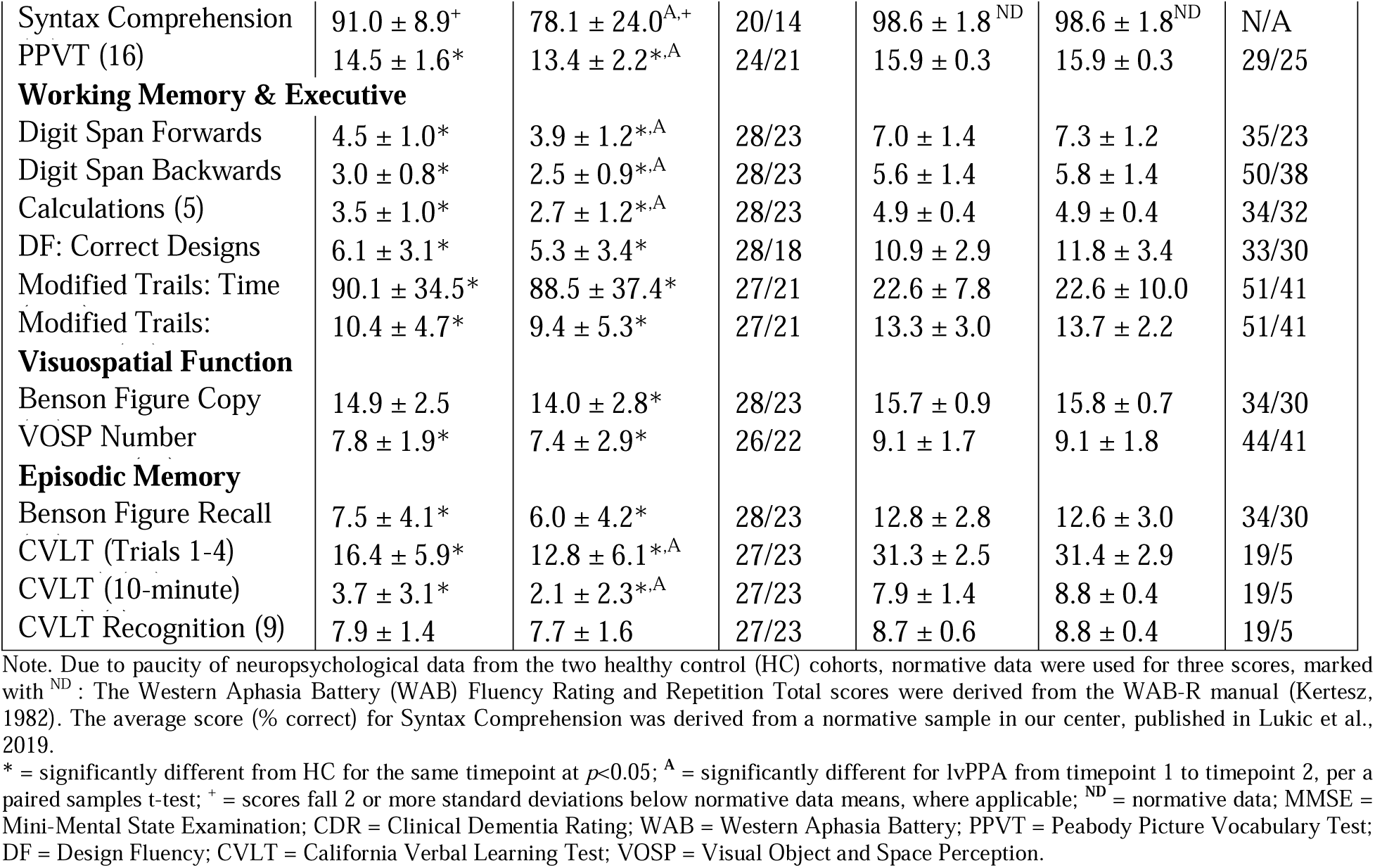
Summary of demographic and cognitive profiles for lvPPA_long_ and HC_long_ cohorts across the timepoints.

### 3.2 Specific anatomical regions in the left temporo-parietal junction showed the greatest cortical atrophy in mild lvPPA

The cross-sectional comparison between the cohort of mild lvPPA (lvPPA_mild_) and the matched group of healthy controls (HC_cross_) yielded the typical asymmetric pattern of atrophy, with the left hemisphere showing more significant involvement than the right. Specifically, significant atrophy was observed in the left inferior parietal lobule and the temporo-parietal junction, as well as the left lateral temporal cortex, dorsolateral prefrontal cortex, and posterior cingulate cortex. Figure 1 illustrates a surface rendering of the atrophy pattern in the mild cohort of lvPPA (n=15), along with the total cohort of lvPPA (n=86) and the baseline atrophy of the longitudinal cohort (n=28) (see Supplementary Table 1 for statistical details). Notably, the left inferior parietal region showed the greatest atrophy across all three lvPPA cohorts, particularly in the left anterior part of the angular gyrus corresponding to the parietal area F, part m (PFm) of the HCP-MMP1.0 parcellation. Significantly and consistently atrophic brain regions were also detected in the left middle frontal gyrus as well as the superior and middle temporal cortices.

**Figure 1.**
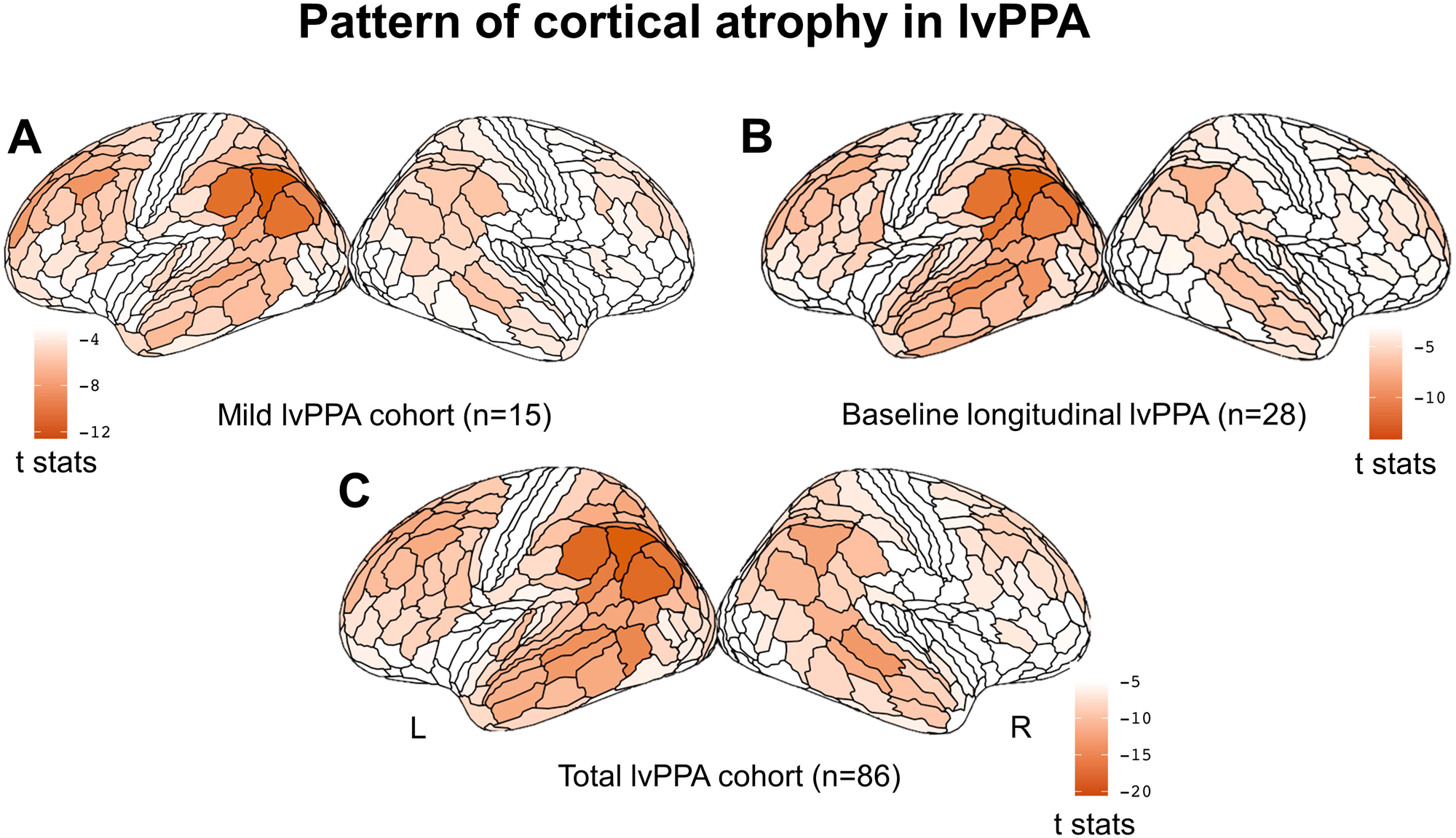
Cross-sectional pattern of cortical atrophy in lvPPA compared to HC. Surface rendering of the atrophy pattern in the mild cohort of lvPPA (n=15) (A), at the baseline atrophy of the longitudinal cohort (n=28) (B), and in the total cohort of lvPPA (n=86). The render is performed with the ggseg toolbox in R (https://lcbc-uio.github.io/ggseg/) on the Glasser brain atlas. The left inferior parietal region showed the greatest atrophy across all three lvPPA cohorts.

In this context, we defined as putative epicenters the regions that were (i) associated with the highest *p*-value and (ii) spatially clustered together within the left parieto-temporal cortex. According to the HCP-MMP1.0 brain parcellation, these regions correspond to the parietal area F, part m (PFm) (coefficient ± SE = -4.28 ± 0.34; *t* = -12.6, *P* < 0.001), to parietal area F (PF) (-3.58 ± 0.31; *t* = -11.5, *P* < 0.001), parietal area G inferior (PGi) (-3.30 ± 0.29; *t* = -11.4, *P* < 0.001), parietal area G superior (PGs) (-3.24 ± 0.32; *t* = -10.2, *P* < 0.001), intraparietal area 0 (IP0) (-2.79 ± 0.29; *t* = -9.7, *P* < 0.001), intraparietal area 2 (IP2) (-3.00 ± 0.32; *t* = -9.5, *P* < 0.001), and Peri-Sylvian Language area (PSL) (-2.71 ± 0.28; *t* = -9.7, *P* < 0.001).

### 3.3 Two partially distinct ICNs were significantly associated with naming and repetition deficits in lvPPA

The seven left hemisphere regions identified in the previous analysis as the earliest and most atrophic regions in lvPPA (PFm, PF, PGi, PGs, lP0, IP2, and PSL) were used as seed-ROIs to extract the corresponding ICNs in the HC_conn_ cohort. In order to ascertain the behavioral relevance of these seven ICNs in relation to the core language deficits in lvPPA, we performed a correlation analysis (involving the full cohort of 86 individuals with lvPPA) between global atrophy (average W-score) within each of the networks (including only the label assigned to the network as indicated in 2.3.4) and the two defining clinical symptoms of lvPPA: impaired confrontation naming and sentence repetition. Due to the non-gaussian distribution of the test scores (BNT W=0.93, p<.001), we used Spearman’s correlation. Given that repetition and naming scores were significantly correlated (Spearman’s *rho* = 0.52, *P* < 0.001), we performed a one-tailed partial correlation analysis (we only expected to observe a positive relationship) between global atrophy (lower values indicate grater global atrophy) and performance (lower scores indicate worse performance) controlling for the influence of the other variable.

Repetition skills (controlling for naming scores) were only significantly correlated with atrophy in the PSL-ICN (*rho* = 0.22, *P* = 0.03), while naming skills (controlling for repetition scores) were only significantly correlated with atrophy in the PFm-ICN (*rho* = 0.21, *P* = 0.03). No other partial correlation reached statistical significance for any of the other networks. In Supplementary Figure 1, we show all the networks identified in the healthy controls and highlight the two networks most relevant to lvPPA symptomatology.

As described in the Methods section, for subsequent analysis, we focused only on the two networks seeded in the left PFm and PSL, which resemble networks previously associated with the typical pattern of atrophy in lvPPA. The PFm network (Fig. 2.A) encompass areas within the inferior parietal and temporal gyri as well as the middle frontal gyrus and resembles the posterior component of the default mode network (DMN). The PSL network (Fig. 2.B) encompass areas within the superior and middle temporal gyri along with the lateral frontal pole and resembles the speech perception network (Battistella et al. 2020). As shown in Figure 2.C, there is some degree of anatomical overlap between these two networks within the perisylvian region, a portion of the inferior temporal lobe and the precuneus. Overall, however, the non-overlapping portions dominate.

**Figure 2.**
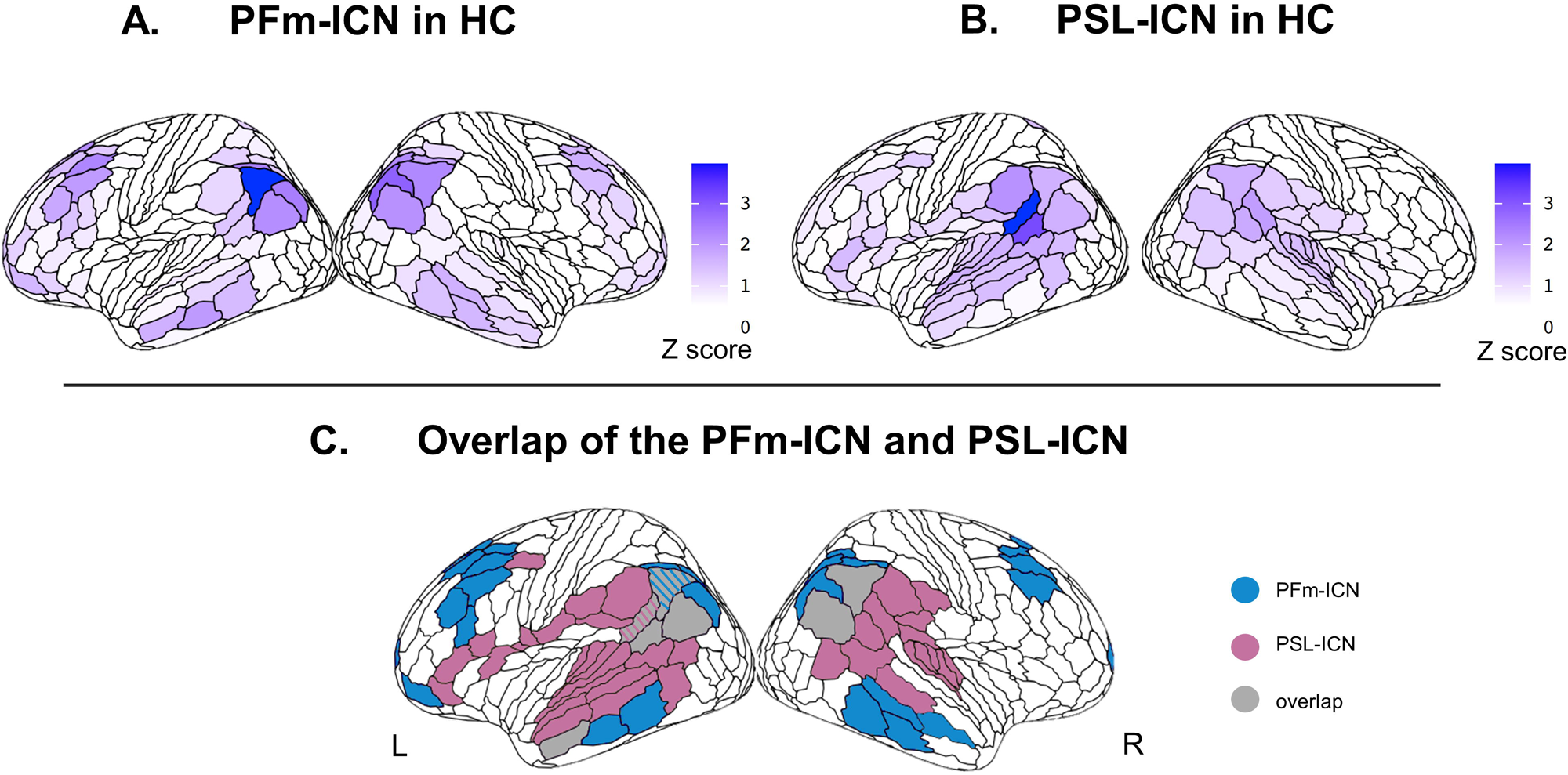
Two partially distinct ICNs associated with naming and repetition deficits in lvPPA. The intrinsic connectivity map (Z score) extracted from the HC_conn_ cohort seeded in the left PFm (A) include areas in the inferior parietal and temporal gyri and in the middle frontal gyrus. The intrinsic connectivity map (Z score) extracted from the HC_conn_ cohort seeded in the left PSL (B) incudes areas in the superior and middle temporal gyri and in the lateral frontal pole. The overlap between these two networks includes regions in the perisylvian region, the inferior temporal lobe and the precuneus (C).

### 3.4 Longitudinal pattern of cortical atrophy in lvPPA

The longitudinal comparison between the lvPPA_long_ and HC_long_ cohorts revealed that the most significant change in cortical thickness occurred bilaterally in the auditory lateral temporal cortices, the inferior parietal cortex (Fig. 3), the posterior cingulate cortex as well as in the left inferior and middle frontal gyri. According to the HCP-MMP1.0 atlas, the regions that showed the greatest change were located in the dorsal portion of the right superior temporal sulcus (STSdp) (coefficient ± SE = -0.043 ± 0.006; *t* = -7.4, *P* < 0.001), followed by the left temporal area F (TF) (-0.044 ± 0.007; *t* = -6.0, *P* < 0.001), the right PGs (-0.043 ± 0.008; *t* = -5.6, *P* < 0.001), the right temporal-parietal-occipital junction area 1 (TPOj1) (-0.028 ± 0.005; *t* = -5.5, *P* < 0.001), the left dorsal 23 section a-b of the posterior cingulate gyrus (d23ab) (-0.035 ± 0.006; *t* = -5.4, *P* < 0.001), the left prostriate cortex (ProS) (-0.048 ± 0.009; *t* = -5.3, *P* < 0.001), the left PGi (-0.034 ± 0.007; *t* = -5.2, *P* < 0.001), and the ventral portion of the left superior temporal sulcus (STSvp) (-0.033 ± 0.007; *t* = -5.0, *P* < 0.001). All the regions along with their statistical significance are provided in Supplementary Table 2.

**Figure 3.**
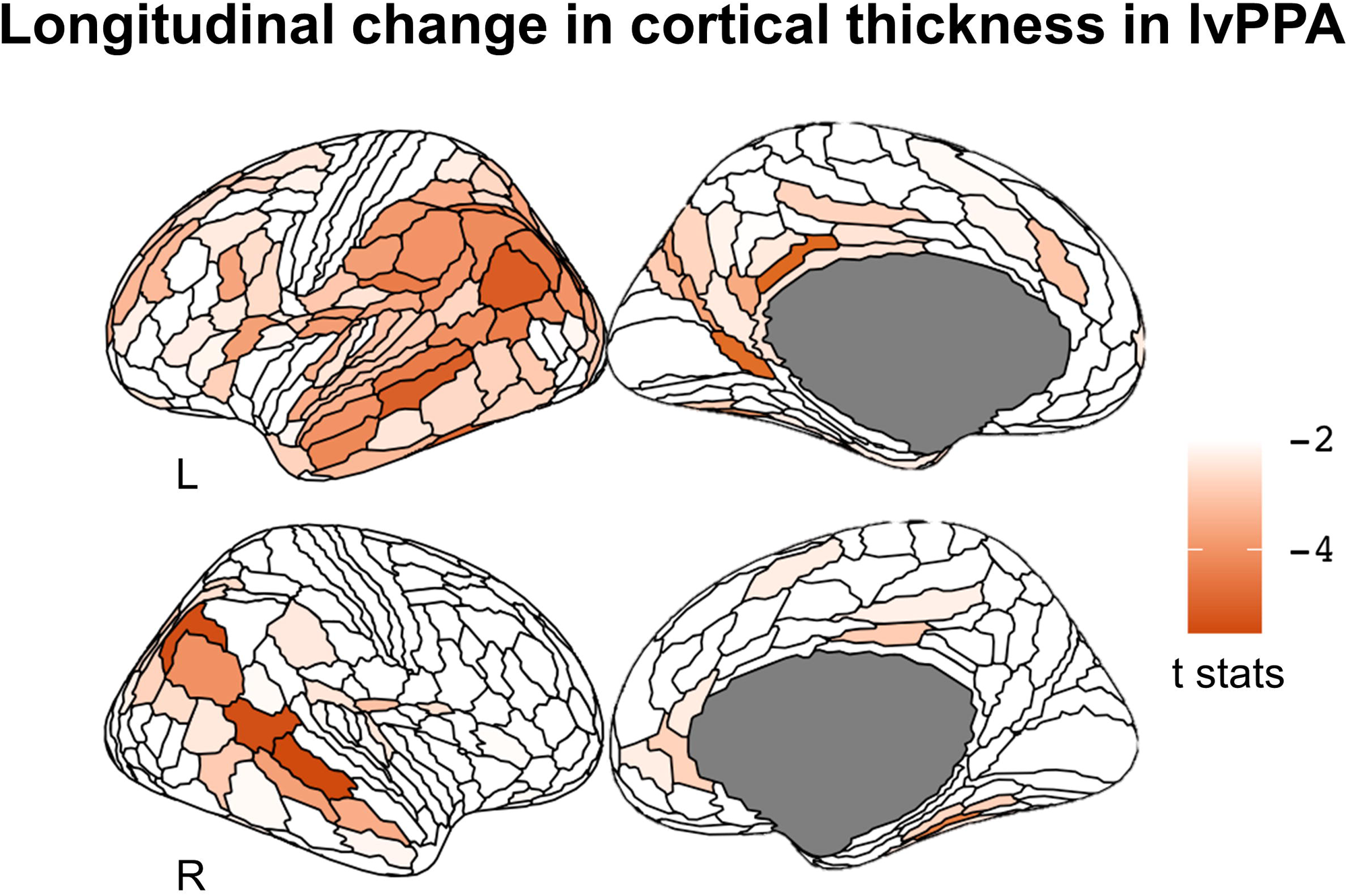
Longitudinal change in cortical thickness in the lvPPA_long_ cohort compared to matched HC with longitudinal data. The longitudinal change in cortical thickness occurred bilaterally in the auditory lateral temporal cortices, the inferior parietal cortex, in the posterior cingulate cortex and in the left inferior and middle frontal gyri. The render is performed with the ggseg toolbox in R (https://lcbc-uio.github.io/ggseg/) on the Glasser brain atlas.

### 3.5 The ICNs associated with sentence repetition and naming deficits were significant predictors of longitudinal atrophy progression in lvPPA

The multiple regression analyses performed between the shortest path length from each ROI to the seed region of each ICN and the ROI-specific t-statistic from the analysis of longitudinal change in cortical thickness in lvPPA (controlling for Euclidian distance) yielded that the two ICNs, which were found to be selectively associated with sentence repetition and naming deficits, were also significant predictors of longitudinal atrophy progression in lvPPA: PFm-ICN (*r* = 0.28, *P* < 0.001) and PSL-ICN (*r* = 0.21, *P* < 0.001). In contrast, longitudinal atrophy progression in lvPPA was not predicted by the shortest path length to the seed region of the salience network (right anterior insula, used as a control here). The corresponding scatterplots for the relationship between the shortest path length to the network epicenters left PFm, left PSL, right AICC and the longitudinal change in cortical thickness (ROI-specific t-statistic) for each ROI of the HCP-MMP1.0 atlas in lvPPA are shown in Fig. 4.

**Figure 4.**
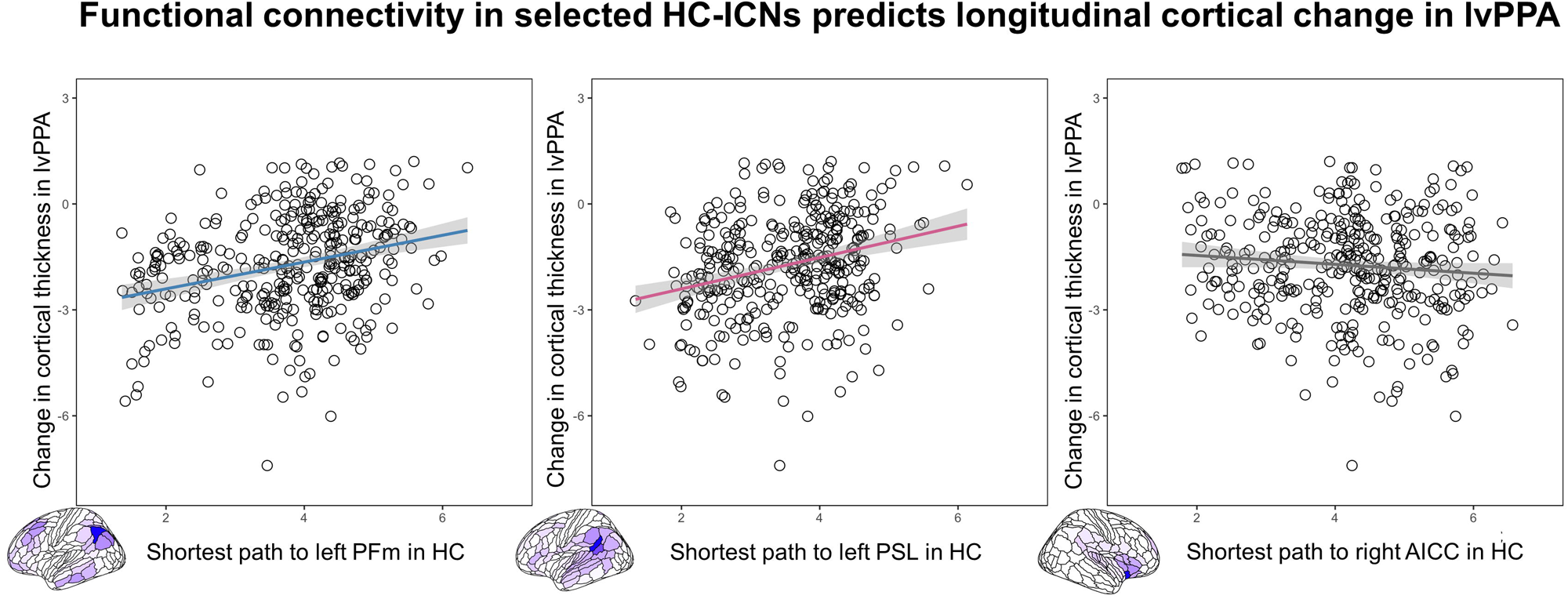
Functional connectivity in selected HC-ICNs predicts longitudinal cortical change in lvPPA. Scatterplots of the correlation between the shortest functional path length from the left PFm, PSL, and right AICC in the HC group and the longitudinal change in cortical thickness in lvPPA. On the scatterplot, each dot represents the statistical change in cortical thickness in each region of the HCP-MMP1.0 atlas.

## 4. Discussion

In this cross-sectional and longitudinal multimodal MR imaging study, we showed that lvPPA aphasic symptomatology and longitudinal atrophy progression were associated with anatomical changes in partly overlapping functional networks anchored to two left parieto-temporal epicenters. These findings have important implication for improving the accuracy with which atrophy epicenters can be localized in diseases with a heterogenous clinical presentation such as lvPPA. Additionally, our results support the model of network-based neurodegeneration, where brain regions connected to disease epicenters are preferentially targeted by neurodegeneration, driving the symptomatology that characterizes each clinical syndrome. These insights can facilitate the development of evidence-based therapies and help to identify anatomically well-defined brain stimulation sites for therapeutic studies. Ultimately, our study contributes to a deeper understanding of the underlying mechanisms of neurodegenerative diseases and the heterogeneity in clinical presentation and progression in lvPPA.

### 4.1 General pattern of atrophy and most vulnerable cortical regions in lvPPA

Consistent with prior studies, we confirmed an asymmetric pattern of atrophy in lvPPA characterized by the cortical involvement of left hemisphere structures including the inferior parietal lobe, the temporo-parietal junction, the dorsolateral prefrontal cortex, the superior and middle temporal gyri and to a lesser extent, regions in the right inferior parietal lobe and in the right middle temporal and frontal gyri (Gorno-Tempini et al. 2004; Mesulam et al. 2009; Henry and Gorno-Tempini 2010; Rogalski et al. 2011; Teichmann et al. 2013; Giannini et al. 2017; Phillips et al. 2018; Preiß et al. 2019). In particular, using a surface-based approach paired with a fine-grained brain parcellation of the cerebral cortex, we localized the most significant atrophy within the left inferior parietal lobe and the posterior-superior tip of the Sylvian fissure at the temporo-parietal junction. While most voxel-based morphometry studies have found atrophy encompassing the middle and superior temporal gyri, it is noteworthy that the most consistently hypometabolic region across PET studies of lvPPA falls in the left inferior parietal lobe (Conca et al. 2022). Interestingly, in our study, the most atrophic peak was located in the left anterior portion of the angular gyrus (PFm), a region that has been described as a transitional area based on its cytoarchitectonic features (Caspers et al. 2006; 2008). The same pattern was observed in the longitudinal lvPPA cohort (using baseline MRI scans) as well as in the larger lvPPA cohort, which spanned a wider range of disease severity. Taken together, these findings indicate that, at the group level, the pattern of brain atrophy is consistent across groups, multifocal, and centered around specific anatomical loci within the left inferior parietal lobe and the temporo-parietal junction (Gorno-Tempini et al. 2011).

Methodological differences might partially explain why accurate identification of early atrophy loci in lvPPA has been problematic. Previous neuroimaging studies typically used standard voxel-based analysis or surface-based approached coupled with coarse brain atlases, preventing precise localization of early atrophic regions in lvPPA. Different nomenclature in and around the inferior parietal regions and temporo-parietal junction may also contribute to the lack of consistency across studies. In this study, we applied a surface-based approach on structural T1-weighted MR images combined with a recent more anatomically-fine-grained brain parcellation atlas that captures the multi-modal fingerprint of each cortical area (Glasser et al. 2016). Critically, compared to voxel-based analyses, surface-based methods have been shown to be most sensitive in capturing atrophy of the cortical ribbon (Diaz-de-Grenu et al. 2014), especially in the parietal lobe, which is characterized by a thinner cortical sheet compared to the temporal lobe (Hogstrom et al. 2013). Indeed, our findings improve neuroanatomical precision by mapping putative atrophy epicenters to a detailed parcellation of cortical areas bounded by sharp changes in architecture, function, connectivity, and/or topography.

### 4.2 Brain networks associated with the core language symptoms of lvPPA

After identifying the specific anatomical location of the most vulnerable regions within the left temporo-parietal cortex in individuals with lvPPA, we used these loci as seed ROIs to delineate their corresponding functional ICNs in healthy individuals and to investigate the behavioral relevance of these networks for the core symptomatology of lvPPA (i.e., impaired sentence repetition and confrontation naming abilities). By employing brain-behavior correlation analyses to evaluate the relation between the structural integrity of each ICN identified in the healthy brain and these core symptoms of lvPPA, we were able to isolate two behaviorally relevant networks: one anchored in the left PFm, which was preferentially associated with auditory-verbal short-term memory (sentence repetition) and one in the left PSL, which was implicated in lexical retrieval (confrontation naming). The two ICNs identified in this study as preferentially involved in sentence repetition and confrontation naming are consistent with findings from previous literature in post-stroke aphasia, functional imaging in healthy controls, and PPA (Fridriksson et al. 2010; Rogalsky et al. 2015; Forkel et al. 2020; Miller et al. 2021). Specifically, language production skills have been linked to a large-scale network involving frontal-temporal-parietal regions (Gleichgerrcht, Fridriksson, and Bonilha 2015; Indefrey and Levelt 2004; Migliaccio et al. 2016; Milton et al. 2021; Mesulam et al. 2014) with the left angular gyrus (including PFm) being specifically associated with impaired confrontation naming (DeLeon et al. 2007) and the temporo-parietal junction (TPJ) and including PSL in repetition (Buchsbaum and D’Esposito 2009; Baldo, Katseff, and Dronkers 2012; Majerus 2013; Lukic et al. 2019).

Interestingly, previous rsfMRI studies were not able to identify a single, one-to-one mapping between the atrophy pattern in lvPPA and a unique functional network related to clinical symptoms and/or disease (Lehmann et al. 2013; Ossenkoppele et al. 2015; Battistella et al. 2020). When considering language-related networks, Battistella et al. 2020 found that lvPPA was the only PPA variant not associated with one specific functional network. Similarly, studies that considered lvPPA and other AD clinical phenotypes (Lehmann et al. 2013; Ossenkoppele et al. 2015) found an overlap between the lvPPA atrophy pattern and the posterior DMN (middle temporal gyrus, inferior parietal lobule and posterior cingulate cortex) as well as “off-DMN” regions (left superior and inferior temporal cortex). Our analysis, by further partitioning the networks previously considered, revealed an association between specific network sub-components and the most relevant clinical symptoms in lvPPA. Therefore, the observations from the present study, in combination with prior evidence, supports the notion that the multifaceted linguistic and cognitive profile of lvPPA might arise from a neurodegenerative process that preferentially targets networks sub-components, potentially explaining the difficulties in defining this variant as a unitary clinical syndrome (Hu et al. 2010; Sajjadi et al. 2012; Machulda et al. 2013; Leyton et al. 2015; Louwersheimer et al. 2016; Ramanan et al. 2020). Future studies might leverage these findings to better understand heterogeneity in the clinical presentation and progression of lvPPA and other atypical AD clinical phenotypes.

### 4.3 Brain networks associated with naming and repetition predict atrophy progression in lvPPA

The results of our longitudinal analysis of cortical thickness changes are in line with previous reports showing marked progression of atrophy over time in lvPPA within functional networks anchored to syndrome-specific, behaviorally relevant epicenters. According to prior studies, atrophy remains asymmetrical in temporo-parietal regions (left > right) despite significant bilateral involvement (Rogalski et al. 2011; Rohrer et al. 2013). In our study, the most prominent longitudinal atrophy change was observed bilaterally in temporo-parietal regions that were also involved in mildly impaired lvPPA patients but to a lesser extent, thus showing the most marked change. Critically, we found that longitudinal spread of atrophy was significantly predicted by the estimated (in healthy controls) length of the shortest functional path to the epicenter of the two networks associated with the two core language symptoms of lvPPA. Informed by the network-based neurodegeneration hypothesis, we anticipated that the greatest cortical thinning over time would occur primarily in the regions that show the strongest functional connectivity with the disease epicenter (the initial peak of atrophy). Indeed, we confirmed that the brain network anchored in the most atrophic region in lvPPA (PFm) was behaviorally relevant (associated with lvPPA symptomatology) and predicted longitudinal atrophy spread. This functional brain network appears to overlap with the DMN that has previously been implicated in AD pathology (Greicius et al. 2004; Buckner 2004). Less expectedly, however, we found a second brain network anchored in a separate temporo-parietal area critical for language that was also involved in the spread of cortical atrophy in lvPPA. Therefore, these findings strongly indicate that more than one predefined large-scale functional system in the brain shapes the pattern of progression of cortical atrophy in lvPPA and drives its associated symptomatology.

Considering that we intentionally chose to select only individuals with lvPPA who had either a positive PIB-PET scan or autopsy-proven AD pathology as part of the longitudinal cohort, it seems sensible to assume that AD is very likely to be the primary underlying neuropathology driving the spread in our patients. In this context, it has been shown that amyloid-beta depositions are typically located in highly inter-connected cortical regions (Buckner et al. 2009; Lehmann et al. 2013), which may facilitate the spread of AD pathology across functionally coupled networks compared to other pathologies that frequently underlie other PPA variants such as FTLD-tau or TDP (Spinelli et al. 2017). Therefore, it could be argued that the multifaceted nature of the core language symptoms in lvPPA with probable AD pathology may arise from the disruption of multiple, distinct functional networks. Although we could not directly investigate the relationship between longitudinal atrophy progression and decline in language abilities due to missing behavioral data, we did observe worsening of naming and repetition skills over time in lvPPA, with a slightly greater effect for naming than repetition which might be explained by the prominent longitudinal thinning of temporal areas bilaterally. Nonetheless, the verbal short term memory deficit in our sample was clearly demonstrated by cross-sectional and longitudinal differences in digit span forward, in addition to WAB sentence repetition, providing additional support to theories that posit phonologic working memory dysfunction as a central feature of AD-associated PPA (Gorno-Tempini et al. 2008; Giannini et al. 2017). More direct evidence comes from a recent study of PPA patients (nfvPPA, svPPA and lvPPA) with autopsy-confirmed AD pathology showing that the rate of decline in naming and repetition skills correlated with the pathological burden (quantified at the time of autopsy) in different left peri-Sylvian regions (Cousins et al. 2021).

### 4.4 Limitations of the study

We acknowledge several limitations of our study. The seed-ROI approach used in this study has the advantage of being more interpretable and straightforward, but global analyses of network modularity are needed to better support any spatial distinction drawn between the two identified networks. While the use of the Euclidean distance for estimating spatial proximity is a widely adopted method, other distance metrics, such as geodesic distance along the cortical sheet or the length of deep white matter tracts, could provide a more accurate reflection of anatomical connectivity between regions. Furthermore, future studies with larger sample sizes or aggregated multicentric datasets, as well as comprehensive profiling of cognitive abilities (both verbal and non-verbal), will be necessary to assesses the generalizability of our findings (including the replicability of putative disease epicenters), to determine the specific language functions subserved by these networks and to explore the potential existence of endophenotypes in lvPPA. Despite these limitations, our study provides valuable insights into the network anatomy of lvPPA and lays the groundwork for future research in this field.

## 5. Conclusion

Taken together, these findings indicate that atrophy progression in lvPPA, starting from inferior parietal and temporo-parietal junction regions, predominantly follows at least two partially distinct pathways, which are associated with auditory-verbal short-term memory and lexical retrieval skills. This indicates that more than one large-scale functional system in the brain influences the pattern of cortical atrophy progression in lvPPA and drives its associated symptomatology. This may help explain the heterogeneity in the clinical presentation and prognosis of individuals with lvPPA. By adopting a network-based perspective of neurodegenerative diseases, we have provided new insights into the network anatomy underpinning lvPPA, therefore advancing scientific knowledge of disease pathogenesis and providing new perspectives to inform future studies investigating the existence of varying endophenotypes in lvPPA.

## Data Availability

While we are able to share anonymized data, public archiving is not yet permitted under the study's IRB approval due to the sensitive nature of the data. Specific requests can be submitted through the UCSF MAC Resource (Request form: http://memory.ucsf.edu/resources/data). Following a UCSF regulated procedure, access will be granted to designated individuals in line with ethical guidelines on the reuse of sensitive data.

## Acknowledgements

The authors thank the participants and their families for the time and effort they dedicated to the research.

## Figure captions

**Supplementary Figure 1. Intrinsic Connectivity Network maps (ICNs) in the healthy control (HC) group and statistical relevance to the lvPPA symptomatology:** Intrinsic connectivity maps (Z score) extracted from the HC_conn_ cohort using the seven atrophic epicenters (PSL, PFm, PF, PGi, PGs, IP2, IP0) identified in the lvPPA_mild_ cohort as seeds. The bar chart represents the Spearman’s coefficient of the partial correlation performed between global atrophy in each network and performance in sentence repetition and confrontation naming controlling for one variable while accounting for the other. The red asterisks indicate the networks in which structural integrity in lvPPA was significantly correlated with repetition (PSL) and confrontation naming (PFm) performance.

