## Supplementary figures and images for "Network anatomy in logopenic variant of primary progressive aphasia"

### Supplemental Figure 1

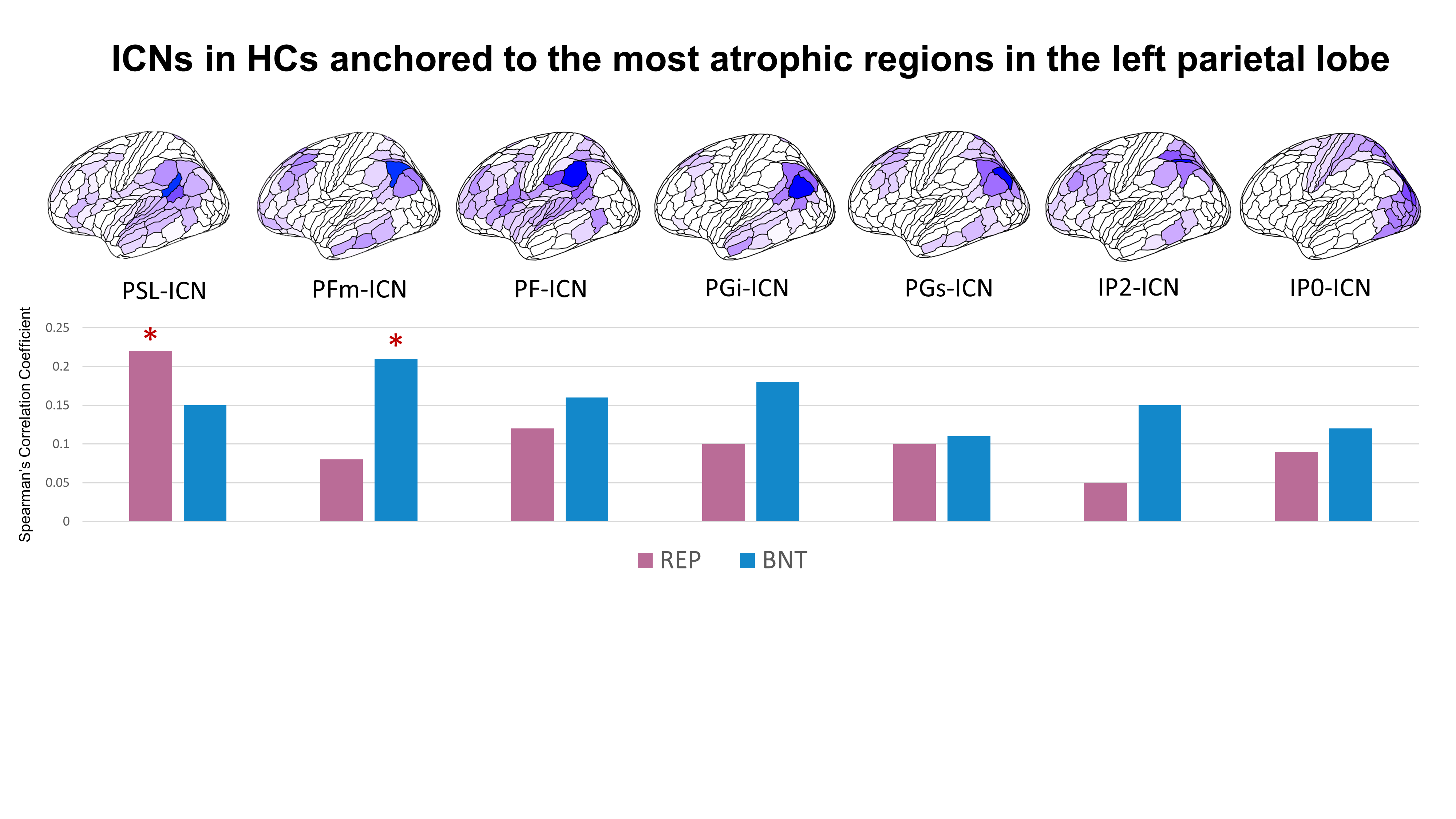
